# Co-development of a best practice checklist for mental health data science: A Delphi study

**DOI:** 10.1101/2021.02.16.21251761

**Authors:** E.J. Kirkham, C.J. Crompton, M.H. Iveson, I. Beange, A. McIntosh, S. Fletcher-Watson

## Abstract

**Background:** Mental health research is commonly affected by difficulties in recruiting and retaining participants, resulting in findings which are based on a sub-sample of those actually living with mental illness. Increasing the use of Big Data for mental health research, especially routinely-collected data, could improve this situation. However, steps to facilitate this must be enacted in collaboration with those who would provide the data - people with mental health conditions.

**Methods:** We used the Delphi method to create a best practice checklist for mental health data science. Twenty participants with both expertise in data science and personal experience of mental illness worked together over three phases. In the Phase 1, participants rated a list of 63 statements and added any statements or topics that were missing. Statements receiving a mean score of 5 or more (out of 7) were retained. These were then combined with the results of a rapid thematic analysis of participants’ comments to produce a 14-item draft checklist, with each item split into two components: best practice now and best practice in the future. In Phase 2, participants indicated whether or not each item should remain in the checklist, and items that scored more than 50% endorsement were retained. In Phase 3 participants rated their satisfaction with the final checklist.

**Results:** The final checklist was made up of 14 “best practice” items, with each item covering best practice now and best practice in the future. At the end of the three phases, 85% of participants were (very) satisfied with the two best practice checklists, with no participants expressing dissatisfaction.

**Conclusions:** Increased stakeholder involvement is essential at every stage of mental health data science. The checklist produced through this work represents the views of people with experience of mental illness, and it is hoped that it will be used to facilitate trustworthy and innovative research which is inclusive of a wider range of individuals.

## Introduction

Data science, in which knowledge is derived from high volume data sets (McIntosh et al., 2016), holds great potential for mental health research (Simon, 2019). Specifically, large quantities of routinely-collected data, such as NHS health records, represent an opportunity to overcome one of the greatest problems previously inherent to such research: recruiting and retaining a representative sample of participants (Furimsky, Cheung, Dewa, & Zipursky, 2008; Woodall, Morgan, Sloan, & Howard, 2010). Recruitment in itself is often time consuming and can be one of the most challenging parts of a research study, whilst the recruitment of a representative sample is harder still (Martin et al., 2018). In mental health research, the representativeness of a sample can be influenced by numerous factors, including clinicians’ willingness to refer participants (Patterson, Kramo, Soteriou, & Crawford, 2010), the severity of participants’ mental illness (Lally et al., 2018), and participants’ employment status (Kannisto et al., 2017). Of particular concern is evidence that people from ethnic minorities are under-represented in mental health research (Gulsuner et al., 2020; Iwamasa, Sorocco, & Koonce, 2002), despite having higher rates of diagnosis for some conditions, such as psychosis (Coid et al., 2008). These factors mean that the groups of individuals who take part in mental health research studies rarely represent the population of people living with mental illness (Kline et al., 2019), which could have serious implications for treatment outcomes (Iltis et al., 2013).

Increased use of routinely-collected mental health data is likely to make research more inclusive and to contribute to the development of more tailored treatments (Iltis et al., 2013; McIntosh et al., 2016). However, work which uses routinely-collected health data, especially sensitive mental health data (King, Brankovic, & Gillard, 2012; Martínez & Farhan, 2019), relies upon the trust of the public whose data are being accessed (Aitken, Jorre, Pagliari, Jepson, & Cunningham-Burley, 2016) – by definition, analysis of routinely-collected data does not involve informed consent from the individuals who provide such data. This means that researchers who work with mental health data must understand and incorporate the views of people with experience of mental illness in their research practice. Such consultation is especially timely in light of the ongoing rapid expansion within mental health data science; after all, it is these individuals whose lives mental health research seeks to improve (Ford et al., 2019).

To this end, we sought to generate a best practice checklist for use in mental health data science to support research that is both rigorous and trusted by those who provide mental health data. The checklist was designed to complement other guidance regarding good practice within data science, such as the UK Data Ethics Framework (Government Digital Service, 2020), the UK Government’s Code of Conduct for Data-Driven Health and Care Technology (Department of Health and Social Care, 2019), and recent work on the development of data governance for the use of clinical free-text data (Jones et al., 2020). Its unique contribution is to encapsulate the perspective of people with lived experience of mental illness, without making recommendations that contravene existing data science frameworks.

To create a checklist that enshrines the principles of trustworthiness and patient-driven priorities within existing research practice, we worked directly with people with expertise in both mental illness and data science. We chose to use the Delphi method, an iterative process in which a group of experts anonymously contributes to the development of consensus on a given topic (Okoli & Pawlowski, 2004). Delphi studies, which typically recruit between 15 and 30 experts (De Villiers, De Villiers, & Kent, 2005), have previously been used to derive guidelines in mental health-related areas such as post-disaster psychosocial care, and first aid recommendations for psychosis and suicidal ideation (Bisson et al., 2010; Jorm, 2015; Kelly, Jorm, Kitchener, & Langlands, 2008; Langlands, Jorm, Kelly, & Kitchener, 2008). Whilst some previous studies have sought to consolidate the views of distinct groups of stakeholders (Murphy, Thorpe, Trefusis, & Kousoulis, 2020), we took the approach of recruiting people with personal or professional expertise in both mental illness and data science. This ensured that the participants themselves were in a position to weigh up the relative merits of the information from both perspectives, and reduced the need for researcher involvement in handling potential trade-offs. The Delphi took place over three phases; each phase involved an online survey completed by participants, followed by analysis by the research team and creation of the next survey (Figure 1).

**Figure 1.**
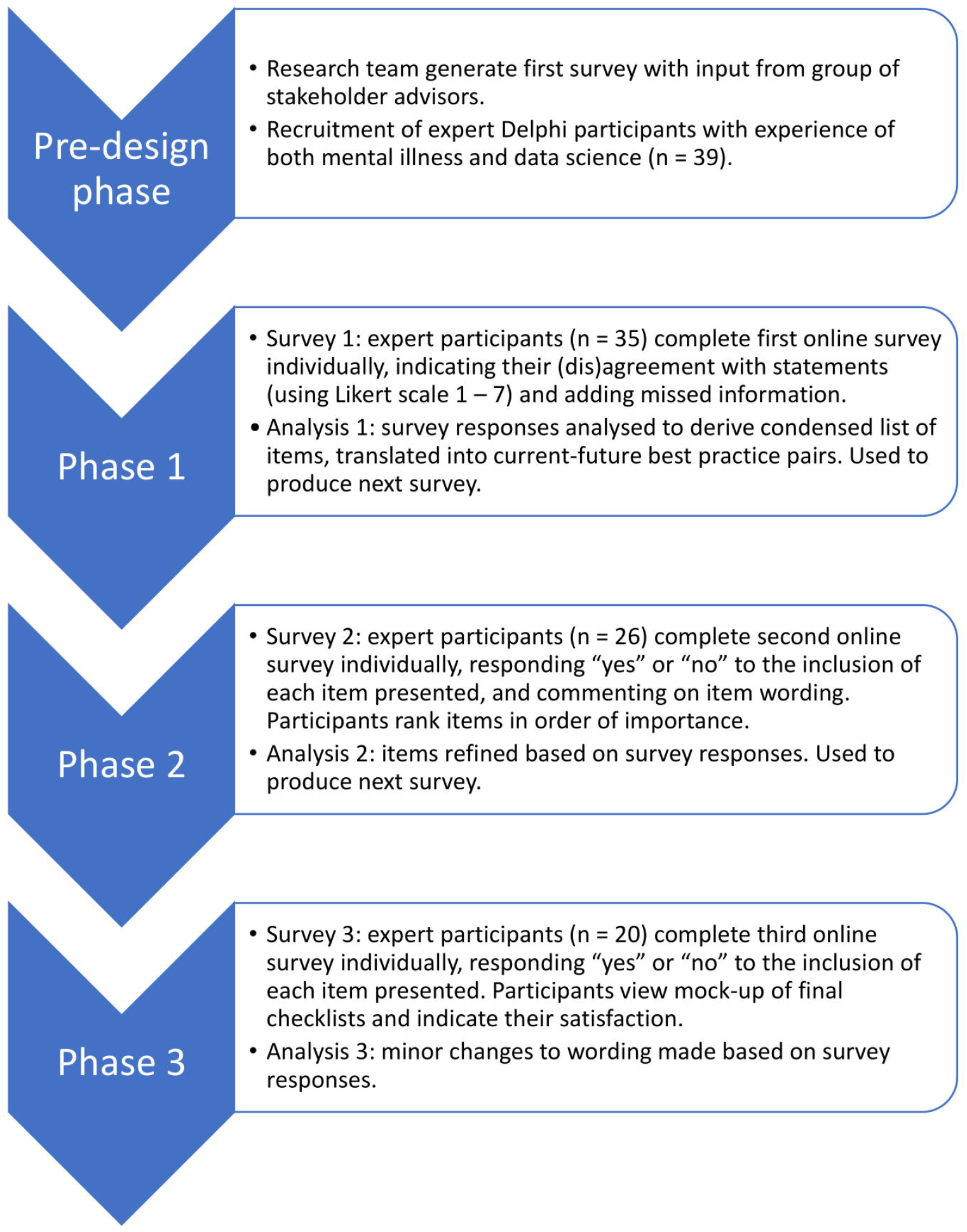
Illustration of the phases of the Delphi process.

## Methods

### Pre-design phase

A pre-design phase was used to identify topics and statements to be included in the first phase of the Delphi study. Initial discussion amongst the research team generated a series of statements which covered the following categories relating to data sharing: users of data, access to data, data linkage, anonymity and de-identification, consent, governance, and community. The research team then produced a draft version of the Phase 1 survey and presented it to a group of people (n = 4) with expertise in mental health, including people with lived experience of mental illness and psychology researchers. These individuals were independent of the participants who took part in the three key phases of the Delphi study. The group recommended improvements primarily based around clarity and usability, and the research team updated the survey accordingly.

### Participants

Participants were recruited if they had both experience with mental illness and experience with data science or research methods. The criteria used to assess this experience, and the number of participants within the final sample who fell into each category, are listed in Table 1 (note that most participants fell under multiple criteria within each category). Before entry into the study potential participants were asked by email to confirm that they met at least one of the criteria in each category. Notably, all participants included in the final sample reported that they had been diagnosed with mental illness at some time in their life, and the majority considered themselves to be living with mental illness when they took part in the study.

**Table 1:** Participants’ experience relevant to mental illness and data science or research methods

| Experience relevant to mental illness | Number | Experience relevant to data science or research methods | Number |
| --- | --- | --- | --- |
| I have been diagnosed with a mental illness at some time in my life. | 20 | I have an undergraduate qualification in an area relevant to data science or research methods<br>(*e.g. psychology, clinical science, epidemiology, statistics etc.*). | 13 |
| I consider myself to have a mental illness at the moment. | 16 | I have a postgraduate qualification in an area relevant to data science or research methods<br>(*e.g. psychology, clinical science, epidemiology, statistics etc.*). | 14 |
| I have family members or close friends who live with mental illness. | 15 | I have advised on a research study/worked on a research team. | 17 |
| I work in an occupation which is related to mental health. | 10 | I work or have worked as a peer researcher. | 6 |
| I am a mental health practitioner. | 2 | I work or have worked in a research setting. | 15 |
|  |  | I work or have worked in a setting that interfaces with research<br>(*e.g. third sector organisation, NHS*). | 14 |

*Note.* Participants included in this table are those who took part in all three phases, n = 20. Most participants fell into more than one category per column.

Participants were recruited through a snowballing technique with relevant contacts and through social media. Thirty-nine participants agreed by email to take part in the study. Of these, 35 individuals provided sufficient data for analysis, though one participant was excluded due to insufficient experience of mental illness and four participants were excluded due to a technical error. Consequently, 30 participants were included in Phase 1 of the study. Of these, 26 provided sufficient data for analysis during Phase 2 of the study, and of these, 20 provided data for Phase 3. Participants who did not provide data for a given phase were excluded from subsequent phases. It should be noted that data collection coincided with the start of the Covid-19 pandemic and UK lock-down, which could have increased drop-out. Nevertheless, a sample of 20 participants is considered a good sample size for a Delphi study (De Villiers et al., 2005).

### Overview of phases

An illustrative overview of the phases included in the Delphi study can be found in Figure 1. The research received ethical approval from the School of Health in Social Science Ethics Committee, University of Edinburgh, ref STAFF212. Potential participants were sent an overview information sheet by email, and those who chose to take part were provided with this overview again at the start of the first survey. All participants provided informed consent to the whole Delphi study at the beginning of the first online survey by responding by tick box to a series of consent statements. An additional, phase-specific information sheet was provided at the beginning of each of the three phases. All data were collected online using surveys hosted by Qualtrics (www.qualtrics.com). In all three surveys, words whose meaning could have been unfamiliar or ambiguous were highlighted in red, and defined in a glossary that was available to download on each page of the survey (Appendix 1).

### Phase 1

#### Materials and procedure

Table 2 provides definitions for the terms used in the subsequent paragraphs. Participants were given 10 days to complete the first survey, and were sent reminder emails during this period. The survey presented participants with a series of 63 statements (Appendix 2). The statements were organised into seven categories (users of data, access to data, data linkage, anonymity and de-identification, consent, governance and community), each of which was divided into two or three sub-categories (Table 3). Each sub-category contained between 2 and 5 statements, and each category (i.e. the combination of sub-categories) contained between 8 and 10 statements. Each category was presented on a separate page of the survey, resulting in between 8 and 10 statements per page. Participants were asked to indicate the extent to which they believed that the given statement represented best practice for mental health data science, using a 7-point Likert scale which ranged from “strongly disagree (1)” to “strongly agree (7)”. At the end of each category of statements, participants were presented with a text box in which they could make comments on the wording of the statements within that category. At the end of the survey, participants were presented with an additional text box in which they could enter any topics or statements concerning best practice in mental health data science that had been missed during the survey.

**Table 2:** Terminology used to describe survey contents

| Term | Use in text | Phase |
| --- | --- | --- |
| statement | Refers to one of the 63 statements included in the Phase 1 survey | 1 |
| category | Refers to the seven categories used to sort the 63 statements in Phase 1 | 1 |
| sub-category | Refers to the sub-sections of the seven categories used in Phase 1 | 1 |
| item | One of the 14 items included in the (draft) checklist in Phases 2 and 3. Each item is divided into two components, a “best practice now” component and a “best practice in the future” component. | 2 and 3 |
| component | Each of the 14 checklist items is divided into two components, a “best practice now” component and a “best practice in the future” component. Therefore, there are 28 components in total within the best practice checklist. | 2 and 3 |

**Table 3:**
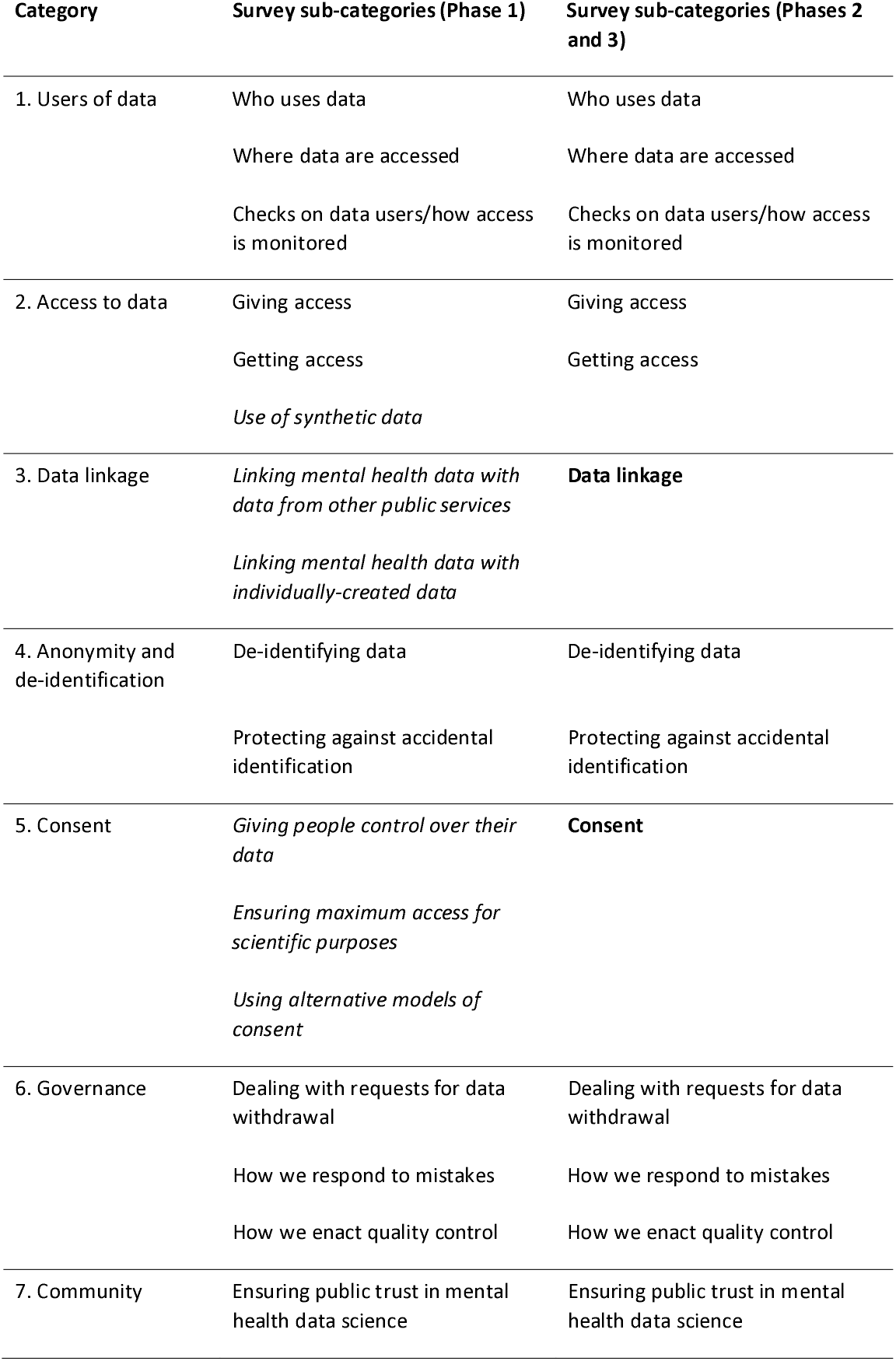
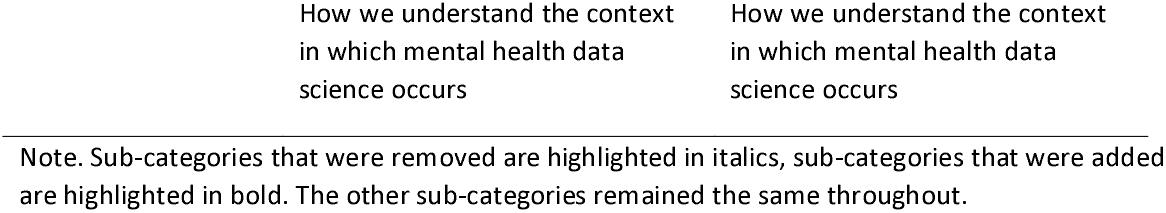
Categories and sub-categories for statements and items included in the surveys

| **Category** | **Survey sub-categories (Phase 1)** | **Survey sub-categories (Phases 2 and 3)** |
| --- | --- | --- |
| 1. Users of data | Who uses data | Who uses data |
|  | Where data are accessed | Where data are accessed |
|  | Checks on data users/how access is monitored | Checks on data users/how access is monitored |
| 2. Access to data | Giving access | Giving access |
|  | Getting access | Getting access |
|  | *Use of synthetic data* |  |
| 3. Data linkage | *Linking mental health data with data from other public services* | **Data linkage** |
|  | *Linking mental health data with individually-created data* |  |
| 4. Anonymity and de-identification | De-identifying data | De-identifying data |
|  | Protecting against accidental identification | Protecting against accidental identification |
| 5. Consent | *Giving people control over their data* | **Consent** |
|  | *Ensuring maximum access for scientific purposes* |  |
|  | *Using alternative models of consent* |  |
| 6. Governance | Dealing with requests for data withdrawal | Dealing with requests for data withdrawal |
|  | How we respond to mistakes | How we respond to mistakes |
|  | How we enact quality control | How we enact quality control |
| 7. Community | Ensuring public trust in mental health data science | Ensuring public trust in mental health data science |

Note. Sub-categories that were removed are highlighted in italics, sub-categories that were added are highlighted in bold. The other sub-categories remained the same throughout.

#### Analysis methods

We began Phase 1 with a large quantity of statements (n = 63, see Appendix 2), some of which contradicted one another, in order to cover as many potential viewpoints as possible. As described above, the original statements were organised by category and sub-category (Table 3). The initial aim of the Phase 1 analysis was to prune and condense the extensive list of statements into one statement per sub-category. The Phase 1 analysis took place over two stages, which we refer to as Stage A and Stage B below (note these “stages” are distinct from the “Phases”, the latter of which represent timeframes of data collection and analysis).

#### Stage A

Stage A involved the initial processing of the quantitative (participant scoring of statements) and qualitative data (participant comments). Statements which received clear support (mean score of more than 5 out of 7) would be retained for Stage B of Phase 1 analysis. Statements which received clear disagreement (mean score of less than 3) were either removed or, alternatively, retained for Stage B but reversed so that they represented the opposite position (Sinclair et al., 2020). Statements with a mean score greater than or equal to 3 and less than or equal to 5 were discarded, due to the absence of a consensus opinion from participants. Next, after quantitative responses to the statements had been analysed, we continued with Stage A by processing the qualitative text responses provided by participants. These data were analysed using a “rapid assessment” version of thematic analysis (McNall & Foster-Fishman, 2007), in which the content of each response was coded and common themes were extracted.

Throughout Stage A analysis, the data were processed within the sub-categories that were present in the Phase 1 survey (Table 3). At the end of Stage A, it was apparent that some of these sub-categories were not useful. For example, quantitative and qualitative responses to the three “consent” sub-categories used in the Phase 1 survey (individual control, scientific access, and alternative models; Table 3) indicated that participants’ views of consent did not fit into these three sub-categories. As a result, a single “consent” category was used instead. In this manner, where appropriate, the sub-categories used in Phase 1 were updated (Table 3).

Once Stage A was complete, we were left with a dataset divided into 14 sub-categories. Each sub-category contained a list of statements that had received clear support (mean score of 5 or more out of 7), and additional recommendations that had been derived from participants’ qualitative responses. Taking for example the sub-category “where data are accessed,” one of the three statements was retained (“…only allowing researchers to see data in specific digital environments (a.k.a. ‘safe havens’)”; see Appendix 2 for statements that were discarded). In addition, the data for this sub-category contained the recommendation, derived from participants’ qualitative responses, that digital controls were preferable to physical controls, a comparison that had not been explicitly addressed within the statements themselves.

#### Stage B

In Stage B of the Phase 1 analysis, all the data for a given sub-category (retained statements and additional information, as described above) were collated. The initial aim of Stage B was to distil all the data within each sub-category into one checklist item. However, when examining the data, it became apparent that in many cases the data collated for each sub-category contained two distinct elements of best practice: recommendations that could be implemented immediately, and those that would depend upon future development and supporting infrastructure. Therefore, instead of following the initial aim of creating one overarching checklist item from the data contained within each sub-category, we decided to create an item which would contain two components: one that referred to best practice that could be implemented immediately, and one that referred to “ideal” best practice to be put in place in future. For example, for the sub-category “responding to mistakes,” the best practice item was divided into a current component: “Best practice for mental health data science means planning in advance to avoid data breaches, utilising a recording process for data breaches, and reporting near misses”, and a future component: “Best practice for mental health data science means developing robust systems to prevent data leaks and breaches”. The newly-created checklist items for each sub-category were then used to create the Phase 2 survey, described below.

### Phase 2

#### Materials and procedure

Participants were given 11 days to complete the second survey, and were sent reminder emails during this period. Participants were presented with the new items (each with a “now” and a “future” component) for each of the 14 sub-categories. For example, one “now” component was “Best practice for mental health data science means allowing other researchers to check analyses wherever possible.” Its corresponding “future” component was “Best practice for mental health data science means providing access to synthetic data where real data cannot be shared, in order to allow other researchers to check analyses.” The full list of items included in this second survey is illustrated in Table 4.

**Table 4:** Phase 2 survey components with their corresponding number of participant comments, divided by comment classification

| Current or future | Best practice for mental health data science means... | Meaningful recommendation | Formatting/language comment | Not relevant/refers to different statement |
| --- | --- | --- | --- | --- |
| Current | Best practice for mental health data science means data should be accessible to a range of people who conduct research, including academics and health workers. | 5 | 2 | 0 |
| Future | Best practice for mental health data science means providing appropriate training and supervision for data users, and carrying out criminal record checks. | 4 | 2 | 0 |
| Current | Best practice for mental health data science means ensuring that data are accessed in safe settings, but that procedures should not be too complicated (to avoid encouragement of unsafe "workarounds"). | 3 | 5 | 0 |
| Future | Best practice for mental health data science means providing digital controls to allow remote access from private settings, using procedures that are not too complicated. | 2 | 5 | 0 |
| Current | Best practice for mental health data science means creating data management plans and ensuring that these are adhered to over time. | 1 | 6 | 0 |
| Future | Best practice for mental health data science means incorporating inspection processes to ensure ongoing compliance with good data practice, and responding proportionately to inappropriate data use with measures such as temporary or long-term suspension of access. | 4 | 3 | 0 |
| Current | Best practice for mental health data science means researchers, scientists and clinical services making data and findings (including null results) open-access where possible, but taking extra care when making decisions regarding access to qualitative data such as free text information. | 6 | 5 | 1 |
| Future | Best practice for mental health data science means building plans for data collected by researchers, scientists and clinical services to be made available for analysis on an open-access basis. | 6 | 1 | 0 |
| Current | Best practice for mental health data science means allowing other researchers to check analyses wherever possible. | 5 | 0 | 0 |
| Future | Best practice for mental health data science means providing access to synthetic data where real data cannot be shared, in order to allow other researchers to check analyses. | 5 | 1 | 0 |
| Current | Best practice for mental health data science means responsible linking of mental health data with other sources of public data, such as education or welfare data, in order to provide new information of public benefit about mental health. | 4 | 2 | 0 |
| Future | Best practice for mental health data science means developing effective measures, including secure linking systems, to protect against identification and misuse. | 3 | 1 | 0 |
| Current | Best practice for mental health data science means using de-identified data, except where identifiable information (including information about protected characteristics) is essential to beneficial outcomes. In all cases the health and benefit of people with lived experience should be prioritised. | 2 | 4 | 0 |
| Future | Best practice for mental health data science means developing methods for de-identification, including innovative ways to mask identifiable information. | 2 | 1 | 0 |
| Current | Best practice for mental health data science means incorporating rules-based statistical disclosure control. | 3 | 7 | 1 |
| Future | Best practice for mental health data science means incorporating principles-based statistical disclosure control with training and external oversight. | 4 | 4 | 1 |
| Current | Best practice for mental health data science means ensuring that participants have as much control over consent as possible. | 4 | 1 | 0 |
| Future | Best practice for mental health data science means exploring alternative models of consent, such as blanket consent for a research topic (e.g. drug development for depression), or blanket consent for a type of data being accessed (e.g. blood test data). | 8 | 1 | 0 |
| Current | Best practice for mental health data science means ensuring that researchers have a process in place for responding to withdrawal requests and that they provide transparency on whether, how and when participants can withdraw their data. | 0 | 0 | 0 |
| Future | Best practice for mental health data science means appointing an independent arbiter to arbitrate on complex questions relating to consent and data withdrawal. | 5 | 0 | 0 |
| Current | Best practice for mental health data science means planning in advance to avoid data breaches, utilising a recording process for data breaches, and reporting near misses. | 1 | 2 | 0 |
| Future | Best practice for mental health data science means developing robust systems to prevent data leaks and breaches. | 5 | 1 | 0 |
| Current | Best practice for mental health data science means monitoring data quality and taking account of the origin and quality of data when drawing conclusions. | 1 | 0 | 0 |
| Future | Best practice for mental health data science means incorporating both stakeholder and procedural oversight of data repositories, with the latter tasked with monitoring data quality and responding to public questions. | 5 | 5 | 0 |
| Current | Best practice for mental health data science means incorporating the views of people with lived experience throughout the course of each project, and providing sensitive and high quality public communication of findings. | 2 | 2 | 0 |
| Future | Best practice for mental health data science means following the principles of open access throughout; publicly pre-registering studies, providing online information of each overarching request to use data and consequent outputs, and publication of null results. | 4 | 3 | 1 |
| Current | Best practice for mental health data science means ensuring that data users understand the underlying data collection tools as well as the socio-cultural context in which studies are designed and findings are disseminated. | 4 | 0 | 0 |
| Future | Best practice for mental health data science means active commitment and working to reduce stigma associated with the phenomena being studied and to increase public understanding of science. | 8 | 1 | 0 |

Note: Number of participants was 26.

For each component, participants were asked to respond “yes” or “no” to whether it should be included in the relevant checklist. After each statement, participants were provided with a text box in which they could make comments on the wording of the component. After this, participants were presented with all the components relating to current best practice (the “now” checklist) and asked to organise them in order of importance. This was then repeated with the future best practice components (the “future” checklist). These rankings are presented in Figures 2 and 3.

**Figure 2.**
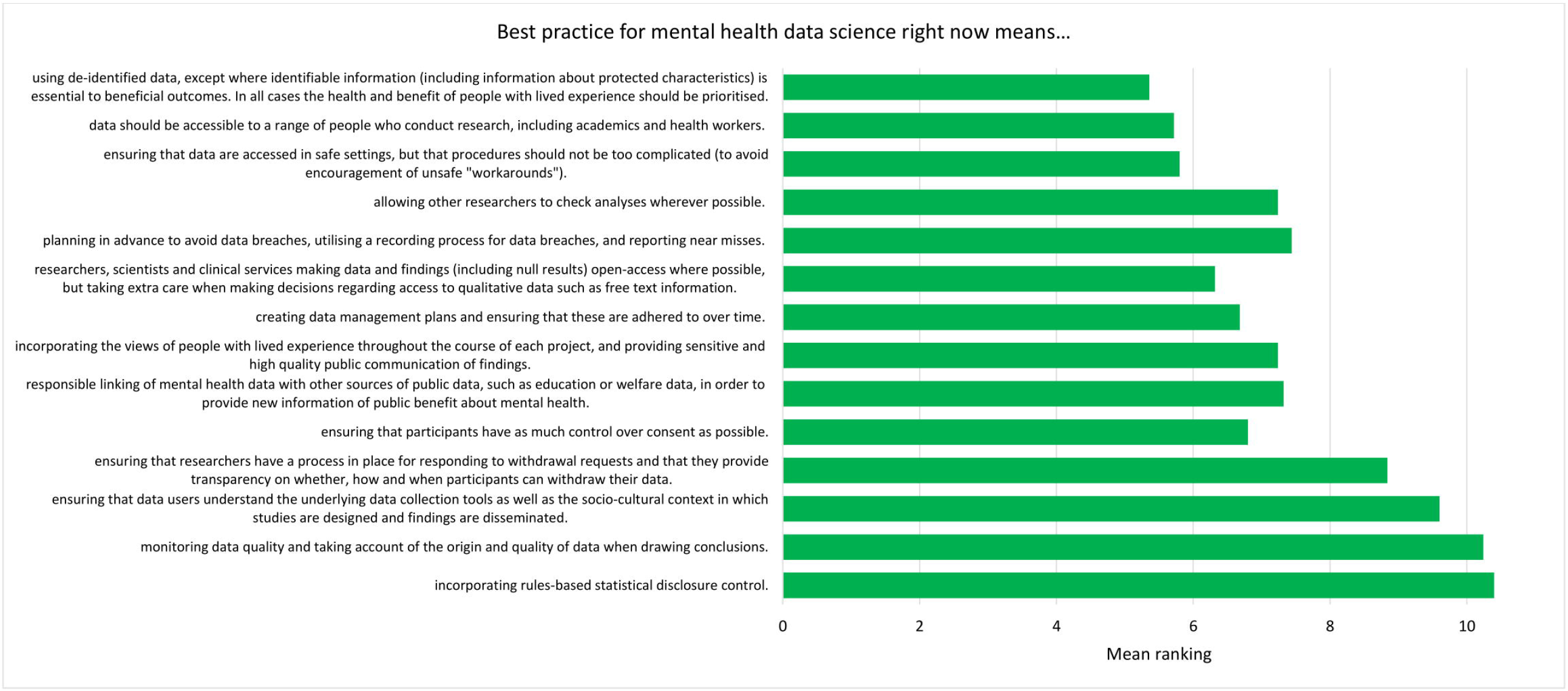
Mean ranking of statements in the current best practice checklist during Phase 2, ordered by median ranking. Lower scores (at the top of the figure) indicate higher importance (number of participants = 25).

**Figure 3.**
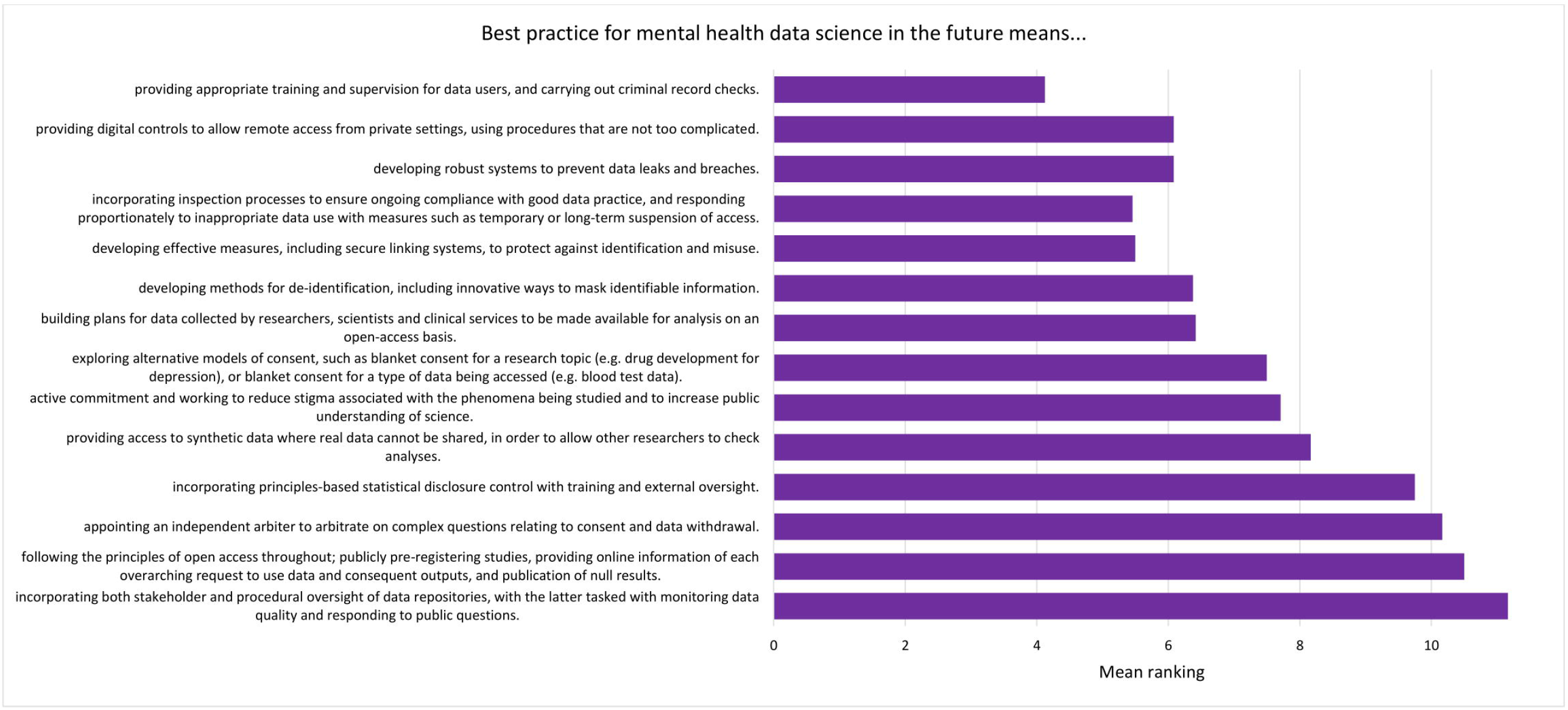
Mean ranking of statements in the future best practice checklist during Phase 2, ordered by median ranking. Lower scores (at the top of the figure) indicate higher importance (number of participants = 24).

#### Analysis methods

The data from the second survey were analysed using both quantitative and qualitative methods. The data used for the quantitative analysis were the yes/no responses provided for each component. Components which received a “yes” response from fewer than 50% of participants were discarded. The qualitative responses, entered in the aforementioned text boxes, were classified into three categories: meaningful recommendation, formatting/language comment, not relevant/refers to different statement. The content of these comments was then used to refine the existing components, and the updated list of components was used to produce the Phase 3 survey.

### Phase 3

#### Materials and procedure

Participants were given 8 days to complete the third survey, and were sent reminder emails during this period. Participants were presented with the updated items derived during the analysis stage of Phase 2. As before, two components (corresponding to current and future best practice) were presented for each of the 14 items. Participants were asked if the given component should be included in the relevant final best practice checklist, and could choose “yes”, “no”, or “other”. If a participant chose “other”, they were presented with a text box in which they could indicate how the component should be changed. Participants were then asked to open a pdf file of a mock-up version of the current best practice checklist, and asked to indicate how satisfied they would be (on a 5-point scale from “very dissatisfied” to “very satisfied”) with a professionally-designed version of this mock-up. This process was repeated for the future best practice checklist.

#### Analysis methods

Statements which received a “yes” response (as opposed to “no” or “other”) from fewer than 50% of participants were discarded. In addition, participants’ comments were assessed and minor changes to the wording of the statements were made. Finally, the median level of satisfaction with each mock-up checklist was measured.

## Results

### Phase 1

#### Stage A

As described in the Methods, the first stage of Phase 1 analysis, Stage A, involved processing the quantitative and qualitative data collected during the Phase 1 survey. Of the 63 original statements, 12 had a mean value between 3 and 5 and were therefore discarded (Appendix 2). The remaining 51 original statements had a mean value of 5 or greater and were retained (Appendix 2). None of the original statements received a mean score of less than 3. Next, participants’ comments were labelled and sorted into themes by EJK, following training from SFW. These themes provided a way of extracting general recommendations from the set of individual participant responses.

#### Stage B

The data (included statements and participant recommendations) were grouped by sub-category. Whilst the initial aim had been to produce one checklist item for each sub-category, it was decided that two components, a “current” and a “future” statement, would be created for each item (see Methods for details). These components are presented in Table 4.

### Phase 2

The list of components included in the Phase 2 survey can be seen in Table 4. All components were given a mean “yes” rating higher than 50%; with the lowest rating at 56%, the highest at 100% and the majority (23/28) falling at or above 75% (see Appendix 3). Therefore, no components were discarded. The number of text responses to each component is presented in Table 4, organised by classification (meaningful recommendation, formatting/language, not relevant/refers to a different statement). These comments were used to update the components for use in Phase 3. Mean and median rankings of component importance were calculated for the “right now” and “in the future” checklists (Figures 2 and 3). Finally, satisfaction ratings for the two checklists were observed (Figure 4). The median satisfaction rating for both checklists was “satisfied”.

**Figure 4.**
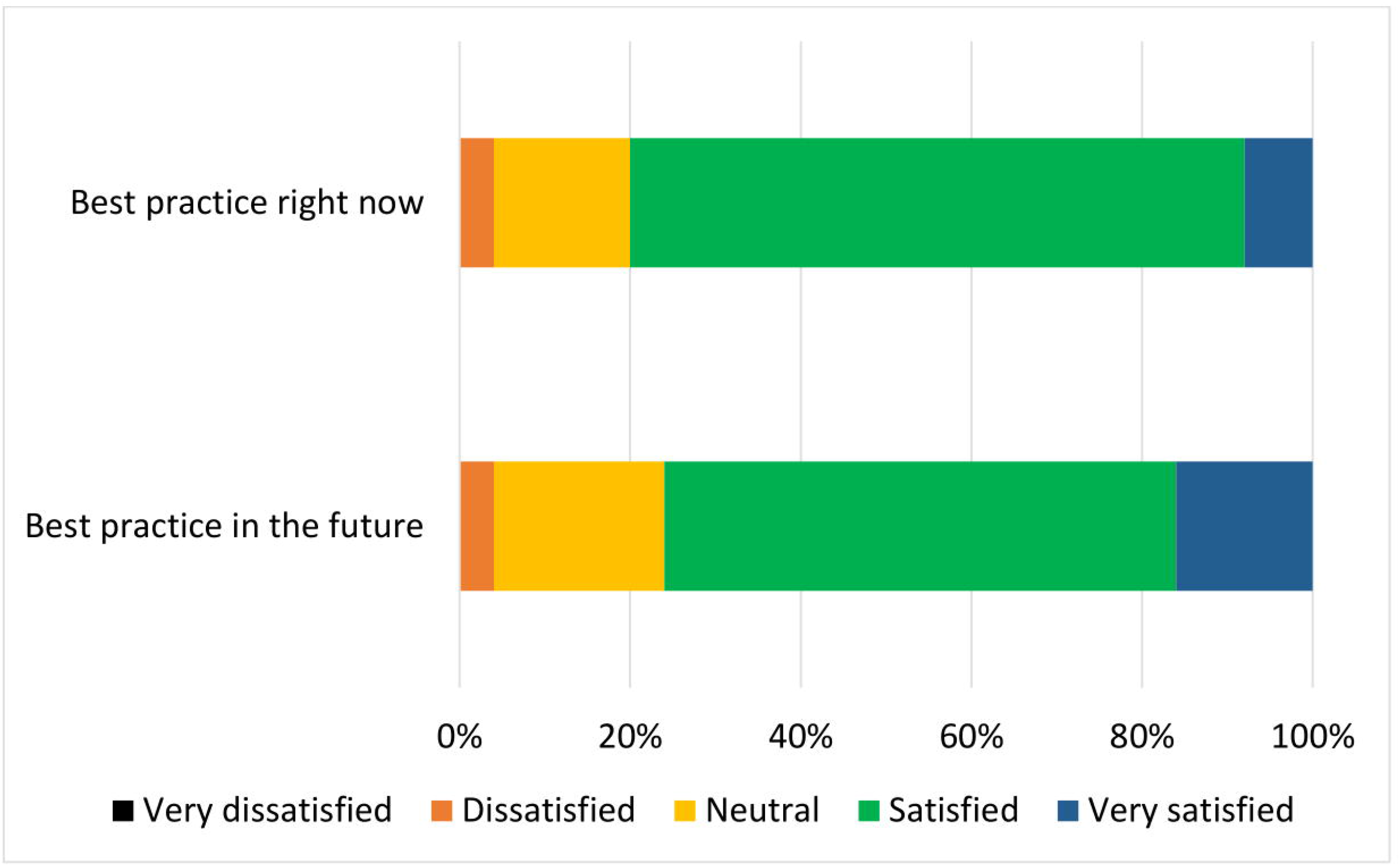
Participant satisfaction with the draft checklist during Phase 2 (n = 25)

### Phase 3

All statements were given a mean “yes” rating higher than 50%; with the lowest rating at 65%, the highest at 100% and the majority (24/28) falling at or above 75% (see Appendix 4). Therefore no statements were discarded. Minor comments regarding the wording of the statements were considered and the statements were altered where appropriate. Satisfaction ratings for the two checklists are illustrated in Figure 5. The median satisfaction rating for both checklists was “satisfied”. The median satisfaction rating for both checklists was “satisfied”. The final versions of the two checklists can be viewed at [OSF link, redacted].

**Figure 5.**
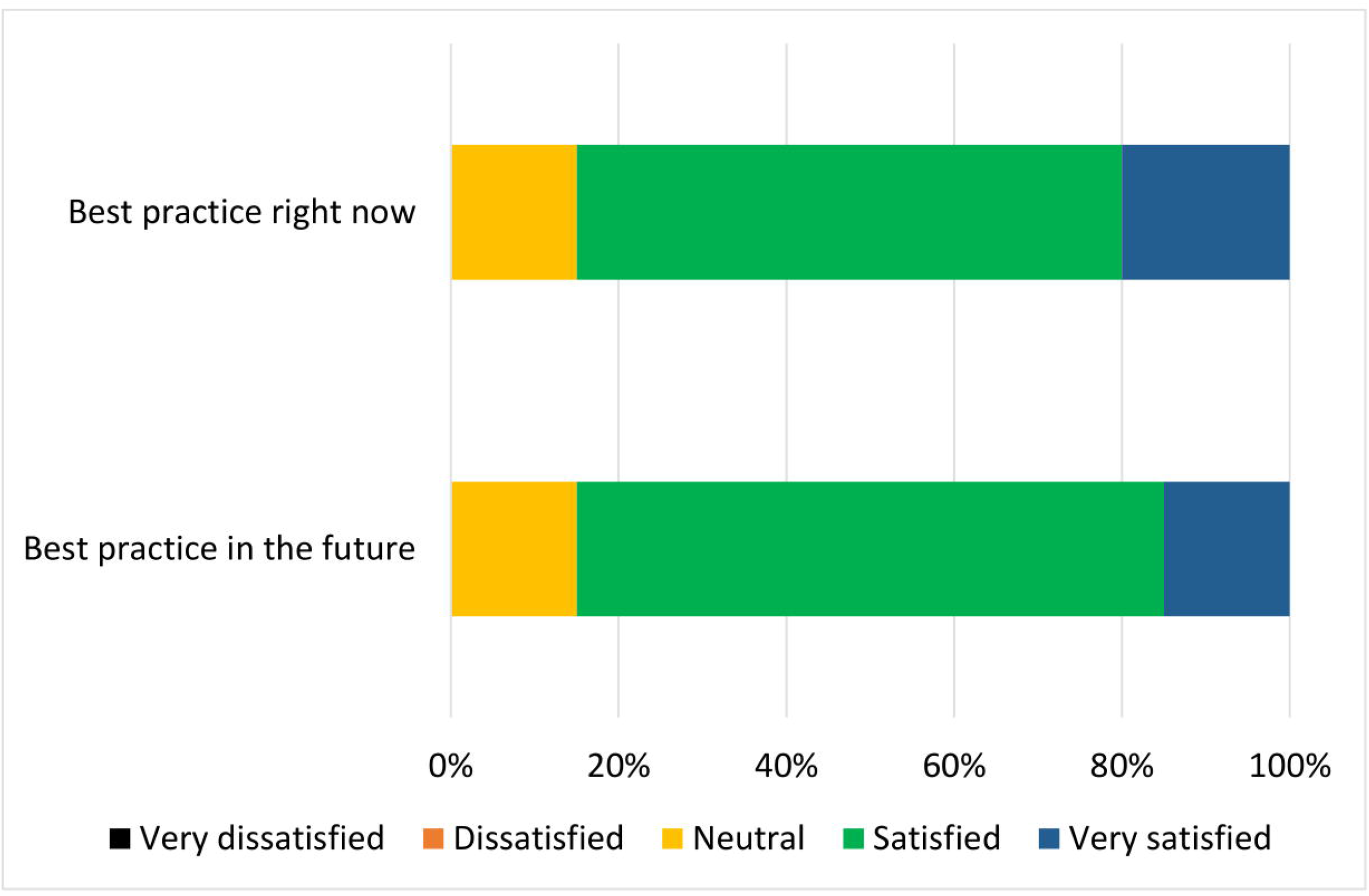
Participant satisfaction with the checklist during Phase 3 (n = 20)

## Discussion

This research used the Delphi method to create guidelines for best practice in mental health data science. A group of participants with expertise in mental illness and data science contributed responses over three phases to produce two checklists: one focused on what mental health researchers can do now, and one focused on what the wider mental health data science community can put in place in the future. Each checklist features 14 items that cover issues pertaining to de-identification of data, data security, transparency and oversight, and community perspectives. Each final checklist was approved by all remaining participants, receiving comprehensive support at the level of individual statements and for the checklist as a whole.

As mental health data science moves forward, it is essential to obtain and maintain the trust of those who provide the data, and represent their views in how data science is conducted (Ford et al., 2019). The use of the Delphi methodology ensures that participants’ involvement goes beyond consultation and becomes part of the scientific process itself. We anticipate that the resultant checklists will be used in conjunction with other work which seeks to apply high standards of data governance to the rapidly evolving field of data science, such as Jones et al.’s (2020) position paper on standards for the use of clinical free-text data.

Examination of the rankings for the current best practice checklist (Figure 2) showed that de-identification of data was expert participants’ highest priority for researchers currently working with mental health data. It is possible that this could reflect particular concerns regarding privacy of mental health data (King et al., 2012), though previous research with the general public suggests that de-identification (which is sometimes perceived as anonymisation; Aitken et al., 2016) is also important when considering health data more generally (Aitken et al., 2016; Buckley, Murphy, & MacFarlane, 2011; Jung, Choi, & Shim, 2020). The rankings also demonstrated an emphasis on keeping data safe and secure, though making data accessible was also viewed as important. This pattern of responses complements previous research with the general public, who were supportive of health data sharing assuming certain conditions (such as trust in those handling the data) were met (Aitken et al., 2016).

The top three ranked statements for the future best practice checklist all fell under the sub-category “users of data”, covering who accesses the data, where it is accessed, and how this access is monitored. The top three statements (Figure 3) reflect a desire for a landscape in which data are well protected via training and supervision for those using the data, and via ongoing oversight of projects. There was a sense that sanctions for inappropriate use of data should be used, but that these should be in proportion to the “offence”; for example, an innocent mistake should be treated differently to deliberate misuse of data, and whole research groups should not be sanctioned based on the behaviour of one individual.

Whilst data protection was clearly of central importance to the expert participants, it became apparent that some current practices for accessing data can be cumbersome and inefficient, especially where physical procedures, such as attending safe settings, are required before access is gained. As one participant noted, this could unintentionally introduce barriers for certain groups of people, such as those with access needs, who may require access to the data. Furthermore, there was concern that extensive and inconvenient barriers to data access could encourage unsafe workarounds, such as using the same password for multiple situations. Delphi participants were generally in favour of moving away from physical controls and towards more efficient, digital control of data. The innovation of more secure, streamlined access to mental health data would have wide-ranging benefits, not least within academia where the time it takes to access data can out-run the length of a project that seeks to study such data (Ford et al., 2019; Iveson & Deary, 2019). This is an important factor for the mental health data science community to consider when designing future procedures and systems.

With respect to the topic of consent, although all participants had expertise in data science, none of the Phase 1 statements concerned with consent procedures which maximise access for scientific research received sufficient consensus for inclusion in later phases (Appendix 2). Consent statements which were retained tended to favour giving individuals control over their data, though a number of individual participants recognised the inherent difficulties with this approach (such as restrictions on research and excessive burden on individuals). Dealing with consent is arguably one of the most challenging issues for the mental health data science community, though innovative models are being developed (Budin-Ljosne et al., 2017; Vayena & Blasimme, 2017). With respect to our research, the expert participants as a group rejected the concept of allowing consent to be provided by a representative sample of participants rather than by all participants, but endorsed the possibility of moving away from individualised models of consent in the future.

With respect to limitations, we acknowledge that there was some drop-out across the three phases, due both to technical error (four participants) and participant attrition (11 participants from Phase 1 to Phase 3). It could be suggested that some of the participants left the study due to disagreement with the concept behind the study. However, this is unlikely given that participants were asked to review an information sheet describing the aims and procedures by sent to them by email before they agreed to begin the study. It is also possible that some of the participant attrition was connected to participants’ mental health, which could have left a final sample who were more well than the sample that initially agreed to take part. Nevertheless, the vast majority of the final sample considered themselves to have a mental illness at the time of the research, suggesting that the views of people with current lived experience were represented. Similarly, we cannot be sure of the extent to which the participants in the present study were representative of people with experience of mental illness more generally; by definition – due to the data science experience criterion - our sample were more highly educated than the general population, and although participants were able to provide additional information about their mental health if they chose to, we did not systematically collect data on the specific mental health conditions they experienced. It is therefore possible that the sample did not represent the full range of people with mental health conditions whose data may be included in future mental health data science research. Finally, whilst a key strength of our study was the inclusion of expert participants with both experience of mental illness and professional knowledge of data science, it is possible that this group of people may differ in their views from those with experience of mental illness but less knowledge of data science. Having said this, given that our study was designed to create guidelines for mental health data science rather than gather opinions on the topic, this is perhaps a less pressing concern.

The underuse of increasingly large sources of data is arguably leading to avoidable health harms (Jones et al., 2017), not least in mental health research, where big data are less widely used than in fields such as oncology or cardiology (McIntosh et al., 2016). However, as mental health data science develops, it is essential that those with experience of mental illness are included every step of the way. Our two resultant checklists focus, respectively, on what mental health researchers working now can do to make their data trustworthy, and on the actions the wider mental health data science community should take in the future. By “in the future”, we refer especially to new platforms for mental health data science; such developments should aim to adhere to the advice provided by the future checklist, and to use it as a guide in the creation of new infrastructure. The rapidly-growing opportunities for using routinely-collected mental health data offer the chance for more inclusive research which captures information from those who are, for whatever reason, unable to engage with traditional research methodologies (Lally et al., 2018; Woodall et al., 2010). We hope that these checklists will facilitate such use, in turn supporting the development of outcomes which include and benefit those who need them most.

## Data Availability

The data is available from the first author and will be made open access in due course.

http://osf.io/9u8ad

## Author contributions

SFW, AM and CC contributed to the conception of the study. EK, SFW, IB, CC and MI contributed to the design of the study. EK and SFW performed the analysis. EK wrote the first draft of the manuscript. CC, SFW, IB and MI made revisions to the draft manuscript. All authors read and approved the submitted version.

## Acknowledgements

The authors would like to thank the Delphi experts who co-produced these guidelines, Mahmud Al-Gailani and VOX Scotland, Suzy Syrett (University of Glasgow), Liz MacWhinney (Lanarkshire Links), the MRC Pathfinder Stakeholder Advisory Group at the University of Edinburgh, our MRC Pathfinder colleagues both within the University of Edinburgh and across the UK, and all those who helped us to recruit our panel of experts.

## Data availability

The datasets used and/or analysed during the current study are available from the corresponding author on reasonable request.

