## Supplementary material for "Co-development of a best practice checklist for mental health data science: A Delphi study"

**Appendix 1: Glossary which accompanied the three surveys**

**Glossary for Delphi study**

**A**

- **academic clinicians:** medical doctors who also carry out research and teaching.
- **anonymous data:** all personal identifying information has been removed and it is no longer possible to trace the information back to the individual. This means if someone found these data, they would not be able to find out who they related to. Unlike with de-identified data, it is impossible link anonymous data to other data relating to the same individual.
- **arbitrate:** when an independent individual makes a judgement on a situation.

C

- **criminal record check:** this is when an employer or organisation applies to the government to see whether an individual has any criminal convictions, cautions, police warnings and so on. There are different levels of check, ranging from “basic” to “enhanced”. In Scotland a criminal record check is sometimes known as a “disclosure check”.

D

- **data breach:** when confidential information is made available outside of a secure environment. A data breach may be intentional or unintentional.
- **data collection tools:** the instruments or devices used to collect data, such as questionnaires or computer programs.
- **data leak:** see data breach.
- **data linkage:** connecting together information (data) – usually at an individual level - from different sources (such as connecting NHS data with school attendance data).
- **data platform:**  a digital location, where data are held for researchers to access and analyse them.
- **data quality checks:** when manual or automated procedures are used to ensure that datasets are complete and accurate.
- **data repository:** a place where multiple datasets from different projects can be stored.
- **data request:** when someone makes a request to have access to a particular type of data (e.g. the ages of everyone who was prescribed drug X in hospital Y).
- **data science:** the use of statistical and computational methods to extract knowledge from large quantities of information (data).
- **data scientists:** people who use statistical and computational methods to extract knowledge from large quantities of information.
- **data type:** this refers to the source of the data. Example data types are blood tests, questionnaires or brain scans.
- **data user:** an individual or organisation who analyses data to find out new knowledge.
- **de-identified data:** data with any information which could be used to identify a person (e.g. name, date of birth) removed.
- **dissemination:** making information available to people outside the research team, through public engagement, reports, publications and so on.
- **duty of candour:** researchers would be expected to be open about their work - to report mistakes and null results, as well as successful, significant findings.

I

- **identifiable information:** any information which can be used to identify a person such as their photograph, their name, their date of birth or their address.
- **independent arbiter:** there are currently systems in place for inspecting some kinds of research (e.g. clinical trials are overseen by the Medicines and Healthcare products Regulatory Agency, MHRA), but there is not yet a dedicated oversight body for data science research. In the case of data science research, an independent arbiter could be a specially trained and trusted supervisor who helps to oversee good data science practice and enforce the rules.
- **informed consent:** when someone gives permission for something to happen whilst being fully aware of the implications and consequences of this.

L

- **lay summary:** a description of a procedure or outcome that can be understood by people without a background in scientific research.

M

- **mental health data:** information that relates to mental health or can be used to learn more about mental health. This could include a range of information, from anti-depressant prescriptions, to weight, to socioeconomic status.
- **mental health data science:** the use of statistical and computational methods to extract knowledge from large quantities of data that relate to mental health.

N

- **null results:** when a researcher does not find a difference between two or more groups being compared.

O

- **open-access online repositories:** digital platforms that hold scientific outputs such as datasets and reports. The information they hold can be accessed by anyone.
- **opt-in procedures:** an individual’s data is not used, unless they consent to their data being used.
- **opt-out procedures:** an individual’s data is used by default, unless they ask for it not to be used.

P

- **peer researcher:** a person with lived experience of the issue being studied who steers and conducts research. This may be in collaboration with other peer researchers and/or researchers without their own lived experience.
- **protected characteristics:** under the Equality Act 2010 it is against the law to discriminate against someone because of their age, disability, gender reassignment, marriage or civil partnership, pregnancy, race, religion or belief, sex or sexual orientation. These are considered protected characteristics.

R

- **representative sample:** a subset of a group which seeks to replicate the characteristics of the whole group. For example, a representative sample may include people from the different age groups and ethnicities that are included in the whole group.
- **research methods:** the processes and techniques that are used to collect evidence for the purpose of generating new or improved knowledge of a topic.
- **routinely-collected data:** data which are originally collected for non-research purposes, such as during the course of NHS care or as part of school administration records.

S

- **safe haven:** secure computing infrastructure in which data are held. Safe havens are access-limited and can only be accessed from accredited locations known as safe settings (see entry for safe settings for more information about these).
- **safe settings:** designated sites located across the UK from which safe havens can be accessed. Access to the physical rooms is controlled and is only allowed to specific approved researchers. Researchers are not allowed to take material in or out, and are often monitored when using these rooms.
- **science communication:** informing non-experts about scientific research and findings.
- **socio-cultural factors**: beliefs, customs and practices which exist within groups of people and influence their thoughts, feelings and behaviours.
- **stakeholder:** an individual who is affected by the processes being carried out. For example, a person with mental illness could be considered a stakeholder in the process of mental health data sharing.
- **statistical disclosure control:** measures applied to data to remove or reduce the risk of individual participants being identified. These measures usually influence the amount of data that can be released.
- **synthetic data:** data which replicates the statistical properties of the original data but changes them in such a way as to remove the possibility of identifying the source of the data.

W

- **withdrawal:** when a participant requests that their data are removed from a dataset after they have taken part in a study. If the results are already published, their data can be removed for new analyses but it will always have been included in the study that was previously published.

**Appendix 2**

*Statements used in the survey during the Phase 1 survey. Scores represent the mean of responses from 30 participants. Statements show in red were discarded at the end of this stage/phase.*

| **Category** | **Sub-category** | **Statement** | **Score** | **Result** |
| --- | --- | --- | --- | --- |
| 1. Users of data | Who uses data | ...acknowledging that the ideal users of data are research scientists including academic clinicians. | 5.37 | Keep |
|  |  | ...ensuring that individual researchers who use data inappropriately are temporarily suspended from accessing data. | 6.30 | Keep |
|  |  | ...ensuring that individual researchers who use data inappropriately are denied future access. | 5.20 | Keep |
|  |  | ...barring a whole research group from using a data platform if one of its researchers is found to have misused data. | 3.07 | Discard |
|  | Where data are accessed | ...preventing researchers from accessing data at home/on their personal computers. | 4.67 | Discard |
|  |  | ...only allowing researchers to see data in specific digital environments (a.k.a. “safe havens”). | 5.17 | Keep |
|  |  | ...only allowing researchers to see data in specific physical locations (a.k.a. “safe settings”). | 4.20 | Discard |
|  | Checks on data users/how access is monitored | ...ensuring that data users have undergone a criminal record check. | 5.30 | Keep |
|  |  | ...making accreditation a prerequisite for access to data relating to protected characteristics. | 5.40 | Keep |
|  |  | ...incorporating inspection processes to ensure compliance with good data practice. | 5.97 | Keep |
| 2. Access to data | Giving access | ...research scientists making their research data accessible to other scientists. | 5.93 | Keep |
|  |  | ...requiring researchers who have collected data to make it available for analysis. | 5.59 | Keep |
|  |  | ...requiring clinical services who hold data to make it available for analysis. | 5.62 | Keep |
|  |  | ...publishing with open-access online repositories (e.g. BioArXiV or the Open Science Framework). | 5.48 | Keep |
|  | Getting access | ...requiring that researchers undergo specific data science training before they can access data. | 6.07 | Keep |
|  |  | ...only allowing data to be accessed by University-based scientists. | 3.59 | Discard |
|  |  | ...removing barriers to data access when health needs are high. | 4.83 | Discard |
|  | Use of synthetic data | ...providing access to synthetic data in cases where real data cannot be shared. | 6.07 | Keep |
|  |  | ...allowing other researchers to check analyses (using synthetic data if necessary). | 6.17 | Keep |
| 3. Data linkage | Linking mental health data with data from other public services | ...maximising benefit by linking data from different sources *e.g. health and education data.* | 5.86 | Keep |
|  |  | ...linking different kinds of routinely-collected data *e.g. NHS data with GCSE results.* | 5.59 | Keep |
|  |  | ...linking education data with mental health data. | 5.83 | Keep |
|  |  | ...linking data from the benefits and welfare system with mental health data. | 5.48 | Keep |
|  |  | ...linking social class data with mental health data. | 5.52 | Keep |
|  | Linking mental health data with individually-created data | ...linking routinely-collected data (e.g. NHS data) with research data (e.g. cohort studies). | 6.14 | Keep |
|  |  | ...linking social media data with mental health data. | 4.52 | Discard |
|  |  | ...linking data from wearable fitness devices (e.g. 'Fitbit') with mental health data. | 5 | Discard |
| 4. Anonymity and de-identification | De-identifying data | ...ensuring that data are de-identified. | 6.38 | Keep |
|  |  | ...removing all data relating to protected characteristics. | 3.72 | Discard |
|  |  | ...removing all identifiable information from datasets (e.g. address, date of birth). | 5.72 | Keep |
|  |  | ...only including data on protected characteristics if this is relevant to the research question. | 6.03 | Keep |
|  |  | ...providing a transparent and clear account of the procedure used to de-identify the data. | 6.86 | Keep |
|  | Protecting against accidental identification | ...incorporating statistical disclosure control. | 6.07 | Keep |
|  |  | ...ensuring that researchers cannot use research data to learn facts about individual people. | 5.79 | Keep |
|  |  | ...placing controls on researchers to ensure data cannot become identifiable through very small subgroups. | 5.93 | Keep |
|  |  | ...placing controls on researchers to ensure data cannot become identifiable through unusual combinations of data. | 5.93 | Keep |
| 5. Consent | Giving people control over their data | ...using opt-in procedures to give control to participants. | 5.61 | Keep |
|  |  | ...ensuring that participants have as much control over consent as possible. | 6.36 | Keep |
|  | Ensuring maximum access for scientific purposes. | ...using opt-out procedures to ensure maximum data usage. | 4.93 | Discard |
|  |  | ...not requiring consent for some types of research. | 3.79 | Discard |
|  |  | ...being aware that consent may not be required if all other best practice measures are in place. | 4.54 | Discard |
|  | Using alternative models of consent | ...tying consent to the research question, *e.g. "I give consent for research relating to drug treatments for depression."* | 5.21 | Keep |
|  |  | ...tying consent to the data type, *e.g. “I give consent for researchers to access my blood test data.”* | 5.64 | Keep |
|  |  | ...allowing consent to be provided by a representative sample of participants rather than by all participants. | 3.61 | Discard |
| 6. Governance | Dealing with requests for data withdrawal | ...providing transparency on the data withdrawal process by providing information on whether, how and when participants can withdraw their data. | 6.75 | Keep |
|  |  | ...ensuring that researchers have a process in place for responding to withdrawal requests. | 6.89 | Keep |
|  |  | ...appointing an independent arbiter to arbitrate on complex questions relating to data withdrawal. | 6.25 | Keep |
|  | How we respond to mistakes | ...utilising a recording process for data breaches and near misses. | 6.79 | Keep |
|  |  | ...planning for what should happen in the case of a data leak or data breach. | 6.86 | Keep |
|  |  | ...applying a duty of candour - to report mistakes and null results, as well as successful, significant findings. | 6.82 | Keep |
|  | How we enact quality control | ...taking account of the origin and quality of the data when drawing conclusions. | 6.86 | Keep |
|  |  | ...incorporating empowered stakeholder oversight of all data repositories. | 5.71 | Keep |
|  |  | ...providing a system for responding to public questions. | 6.39 | Keep |
|  |  | ...incorporating data quality checks. | 6.82 | Keep |
| 7. Community. | Ensuring public trust in mental health data science | ...providing accessible public education on how data are used. | 6.46 | Keep |
|  |  | ...data users building trust with a community of representatives. | 6.39 | Keep |
|  |  | ...acknowledging that data scientists have a responsibility to provide open and public explanations of what they are doing. | 6.54 | Keep |
|  |  | ...providing online information that details each new data request and consequent output. | 6.00 | Keep |
|  |  | ...taking account of socio-cultural factors during the design of studies. | 6.29 | Keep |
|  | How we understand the context in which mental health data science occurs. | ...taking account of socio-cultural factors during the dissemination of findings. | 6.21 | Keep |
|  |  | ...recognising stigma associated with phenomena being studied. | 6.61 | Keep |
|  |  | ...focusing on health and benefit for patients and their families. | 6.43 | Keep |
|  |  | ...ensuring that data users understand the underlying data collection tools used to gather the data. | 6.32 | Keep |

**Appendix 3**

*Statements used in the Phase 2 survey. Scores represent the percentage of “yes” responses from 26 participants. No items were discarded at this stage.*

| **Category** | **Sub-category** | **Checklist** | **Statement (“Best practice for mental health data science means…”)** | **Percent “yes”** | **Result** |
| --- | --- | --- | --- | --- | --- |
| 1. Users of data | Who accesses data | Current | data should be accessible to a range of people who conduct research, including academics and health workers. | 96.15 | Keep |
|  |  | Future | providing appropriate training and supervision for data users, and carrying out criminal record checks. | 92.31 | Keep |
|  | Where users access data | Current | ensuring that data are accessed in safe settings, but that procedures should not be too complicated (to avoid encouragement of unsafe "workarounds"). | 73.08 | Keep |
|  |  | Future | providing digital controls to allow remote access from private settings, using procedures that are not too complicated. | 88.46 | Keep |
|  | How access is monitored | Current | creating data management plans and ensuring that these are adhered to over time. | 96.15 | Keep |
|  |  | Future | incorporating inspection processes to ensure ongoing compliance with good data practice, and responding proportionately to inappropriate data use with measures such as temporary or long-term suspension of access. | 80.77 | Keep |
| 2. Access to data | Giving access to data | Current | researchers, scientists and clinical services making data and findings (including null results) open-access where possible, but taking extra care when making decisions regarding access to qualitative data such as free text information. | 92.31 | Keep |
|  |  | Future | building plans for data collected by researchers, scientists and clinical services to be made available for analysis on an open-access basis. | 84.62 | Keep |
|  | Getting access to data | Current | allowing other researchers to check analyses wherever possible. | 84.62 | Keep |
|  |  | Future | providing access to synthetic data where real data cannot be shared, in order to allow other researchers to check analyses. | 73.08 | Keep |
| 3. Data linkage | Data linkage | Current | responsible linking of mental health data with other sources of public data, such as education or welfare data, in order to provide new information of public benefit about mental health. | 84.62 | Keep |
|  |  | Future | developing effective measures, including secure linking systems, to protect against identification and misuse. | 88.00 | Keep |
| 4. Anonymity and de-identification | How we de-identify data | Current | using de-identified data, except where identifiable information (including information about protected characteristics) is essential to beneficial outcomes. In all cases the health and benefit of people with lived experience should be prioritised. | 92.00 | Keep |
|  |  | Future | developing methods for de-identification, including innovative ways to mask identifiable information. | 92.00 | Keep |
|  | How we protect against accidental identification | Current | incorporating rules-based statistical disclosure control. | 56.00 | Keep |
|  |  | Future | incorporating principles-based statistical disclosure control with training and external oversight. | 84.00 | Keep |
| 5. Consent | Consent | Current | ensuring that participants have as much control over consent as possible. | 80.00 | Keep |
|  |  | Future | exploring alternative models of consent, such as blanket consent for a research topic (e.g. drug development for depression), or blanket consent for a type of data being accessed (e.g. blood test data). | 80.00 | Keep |
| 6. Governance | Dealing with requests for data withdrawal | Current | ensuring that researchers have a process in place for responding to withdrawal requests and that they provide transparency on whether, how and when participants can withdraw their data. | 96.00 | Keep |
|  |  | Future | appointing an independent arbiter to arbitrate on complex questions relating to consent and data withdrawal. | 72.00 | Keep |
|  | Responding to mistakes | Current | planning in advance to avoid data breaches, utilising a recording process for data breaches, and reporting near misses. | 96.00 | Keep |
|  |  | Future | developing robust systems to prevent data leaks and breaches. | 92.00 | Keep |
|  | Enacting quality control | Current | monitoring data quality and taking account of the origin and quality of data when drawing conclusions. | 96.00 | Keep |
|  |  | Future | incorporating both stakeholder and procedural oversight of data repositories, with the latter tasked with monitoring data quality and responding to public questions. | 73.91 | Keep |
| 7. Community | Ensuring public trust in mental health data science | Current | incorporating the views of people with lived experience throughout the course of each project, and providing sensitive and high quality public communication of findings. | 100.00 | Keep |
|  |  | Future | following the principles of open access throughout; publicly pre-registering studies, providing online information of each overarching request to use data and consequent outputs, and publication of null results. | 92.00 | Keep |
|  | Addressing the context in which mental health data science occurs | Current | ensuring that data users understand the underlying data collection tools as well as the socio-cultural context in which studies are designed and findings are disseminated. | 88.00 | Keep |
|  |  | Future | active commitment and working to reduce stigma associated with the phenomena being studied and to increase public understanding of science. | 84.00 | Keep |

**Appendix 4**

*Statements used in the survey during the third phase*

| **Category** | **Sub-category** | **Checklist** | **Statement (“Best practice for mental health data science means…”)** | **Percent “yes”** | **Result** |
| --- | --- | --- | --- | --- | --- |
| 1. Users of data | Who accesses data | Current | data should be accessible to a range of people who  conduct research, including authorised academics and  clinicians. | 100 | Keep |
|  |  | Future | providing appropriate training and supervision for data  users, and carrying out criminal record checks where  relevant. | 75 | Keep |
|  | Where users access data | Current | ensuring that data are accessed in safe settings using  clear and efficient procedures. | 85 | Keep |
|  |  | Future | providing digital controls to allow remote access from  private settings, using procedures that are robust and  easy to follow. | 90 | Keep |
|  | How access is monitored | Current | creating data management plans and ensuring that  these are adhered to at all times. | 95 | Keep |
|  |  | Future | incorporating inspection processes to ensure ongoing  compliance with good data practice, and responding  proportionately to inappropriate data use with  measures such as training or temporary or long-term  suspension of access. | 85 | Keep |
| 2. Access to data | Giving access to data | Current | researchers, scientists and clinical services making deidentified  data and findings (including null results) open access  where possible. It also involves awareness of the  risk that qualitative data (such as free text) could  contain identifiable information. | 80 | Keep |
|  |  | Future | building plans for de-identified data collected by  researchers, scientists and clinical services to be made  available for analysis on an open-access basis. | 80 | Keep |
|  | Getting access to data | Current | allowing other researchers to check analyses wherever  possible (in addition to peer review). | 80 | Keep |
|  |  | Future | providing access to synthetic data where real data  cannot be shared, in order to allow other researchers to  check analyses and conclusions. | 85 | Keep |
| 3. Data linkage | Data linkage | Current | facilitating research linking mental health data with  other sources of public data, such as education or  welfare data, in order to provide new information of  public benefit about mental health. | 80 | Keep |
|  |  | Future | developing effective measures, including secure linking systems, to protect against inappropriate identification  and misuse. | 75 | Keep |
| 4. Anonymity and de-identification | How we de-identify data | Current | using de-identified data, except where identifiable  information (including information about protected  characteristics) is essential to beneficial outcomes. In all  cases the health and benefit of people with lived  experience should be prioritised. | 65 | Keep |
|  |  | Future | developing methods for de-identification, including  innovative ways to mask identifiable information. | 70 | Keep |
|  | How we protect against accidental identification | Current | incorporating statistical disclosure control by following  rules designed to prevent identification of individuals. | 90 | Keep |
|  |  | Future | incorporating statistical disclosure control based on principles, such as the principle that no individual may  be identified, with training and external oversight. | 75 | Keep |
| 5. Consent | Consent | Current | ensuring that participants have as much control over  consent as possible. | 75 | Keep |
|  |  | Future | exploring alternative models of consent, which may  involve moving away from individualised models of  consent. | 75 | Keep |
| 6. Governance | Dealing with requests for data withdrawal | Current | ensuring that researchers have a process in place for  responding to withdrawal requests and that they  provide transparency on whether, how and when participants can withdraw their data. | 95 | Keep |
|  |  | Future | appointing a qualified, independent arbiter to arbitrate  on complex questions relating to consent and data  withdrawal. | 65 | Keep |
|  | Responding to mistakes | Current | planning in advance to prevent data breaches, using a  recording process for data breaches, and reporting  near misses. | 95 | Keep |
|  |  | Future | developing robust systems to prevent data leaks and  record data breaches and near misses. | 80 | Keep |
|  | Enacting quality control | Current | monitoring data quality and taking account of the origin  and quality of data when drawing conclusions. | 85 | Keep |
|  |  | Future | incorporating oversight of data repositories in order to  monitor data quality and respond to public enquiries. | 80 | Keep |
| 7. Community | Ensuring public trust in mental health data science | Current | incorporating the views of people with lived experience  throughout the course of each project, and providing  nuanced and high-quality public communication of  findings. | 80 | Keep |
|  |  | Future | incorporating the views of people with lived experience  throughout the course of each project, whilst following  the principles of open access by publicly pre-registering  studies and providing accessible online information of  each overarching request to use data, each output, and  any null results. | 70 | Keep |
|  | Addressing the context in which mental health data science occurs | Current | ensuring that data users understand the underlying  data collection tools as well as the socio-cultural  context in which studies are designed and findings are  disseminated. | 80 | Keep |
|  |  | Future | active commitment to reducing the stigma associated  with mental illness and its research, and to increasing  public understanding of science. | 95 | Keep |
